# Methylation Risk Score Identifies an Interferon-Driven SLE Subset Distinct from Polygenic Risk

**DOI:** 10.64898/2026.03.10.26348007

**Authors:** Holme Vestin, Nina Oparina, Maija-Leena Eloranta, Elisabeth Skoglund, Ioanna Giannakou, Martina Frodlund, Iva Gunnarsson, Christopher Sjöwall, Elisabet Svenungsson, Lars Rönnblom, Juliana Imgenberg-Kreuz, Dag Leonard

## Abstract

**Objectives:** The aetiopathogenesis of SLE encompasses genetic, environmental and epigenetic factors. We investigated associations between an SLE methylation risk score (MRS), HLA-DRB1*03:01, a non-HLA polygenic risk score (PRS) and clinical and immunological phenotypes.

**Methods:** DNA methylation in whole blood from patients fulfilling ≥4 ACR-82 criteria and controls were investigated using the Illumina HM450K array. The discovery cohort included 311 patients and 400 controls, and the replication cohort comprised 175 patients and 187 controls. Seventeen independent, top differentially methylated CpG sites (Δβ of ≥0.1) from case-control comparisons, were used to calculate the MRS. Genotyping was performed using the Immunochip, and the PRS included 57 non-HLA SLE SNVs. Clinical data were collected from patient charts, and serum IFN-α2 was measured using Simoa.

**Results:** Higher MRS was strongly associated with serum IFN-α2 levels (p=1.04×10^−14^). In both cohorts, higher MRS associated with discoid lupus, immunologic involvement, and anti-SSA/SSB/RNP/Sm autoantibodies (all p<0.05), and with higher disease activity in the discovery cohort (p=1.50×10⁻⁴). MRS was also elevated in patients with multiple autoantibodies (p<1.0×10^-15^) and in HLA-DRB1*03:01 carriers (p<1.0×10^-3^). In contrast, higher PRS was associated with nephritis, anti-dsDNA positivity, and lower prevalence of anti-SSB antibodies (all p<0.05). No correlation was observed between the MRS and the PRS (p=0.35).

**Conclusion:** The MRS defines an interferon-high, HLA-DRB1*03:01-linked SLE subset with multiple autoantibodies, partly distinct from PRS-associated nephritis risk, highlighting potentially divergent pathogenic pathways. These findings underscore the value of integrating genetic and epigenetic data to better understand underlying disease mechanisms in SLE.

**Key Messages:**

- Higher MRS, but not PRS, correlated with increased levels of serum IFN-α.
- The MRS was associated with discoid rash, hematologic disorder, hypocomplementemia, antibodies including anti-SSA and HLA-DRB1*03:01.
- Higher PRS was linked to nephritis and anti-dsDNA positivity, and did not associate with the MRS.

## Introduction

Systemic lupus erythematosus (SLE) is an autoimmune, heterogeneously presenting disease characterised by dysregulated innate and adaptive immune responses (1). The disease is polygenic, although there are monogenic variants that contribute to disease in rare cases (2). So far, 330 genomic susceptibility loci associated with SLE have been identified (3, 4) and a high polygenic risk score (PRS) has been demonstrated to predict more severe disease phenotypes (5). The concordance rate between monozygotic twins has been reported to be 24-69% (6) and the heritability has been estimated to 44% (7).

In addition to genetic status, epigenetic modifications together with environmental exposures including hormonal factors act in a complex interplay ultimately precipitating the immunological aberrations observed in SLE (8). Epigenetic changes principally include DNA methylation, histone modifications and non-coding RNAs (9). DNA methylation occurs when a methyl group is added to the 5’ carbon on the cytosine residue in CpG sites which are dinucleotides composed of cytosine, phosphate and guanine in a 5’ to 3’ direction. This covalent modification typically causes transcriptional silencing through inhibition of transcription factor binding (10).

DNA methylation is subjected to genetic control. In the same cohort as the present study, Imgenberg-Kreuz *et al.* reported genetic regulation in the form of cis-methylation quantitative trait loci (meQTL) at 466 (16%) differentially methylated CpG sites (DMCs) in patients with SLE (11). Further, it was found that 7254 out of 385 851 CpG sites included in the Illumina HM450K array were differentially methylated in patients with SLE, with 75% being hypomethylated and the largest effects being observed at interferon (IFN)-regulated genes.

Interestingly, IFN-α, a type I IFN, has been shown to induce expression of HLA-DRB1 in lupus T cells (12). The HLA-DRB1*03:01 risk allele codes for an epitope that, in the presence of IFN-γ, has been reported to elicit transcriptomic programs that are characteristic for SLE and to alter cellular responses in an SLE-like manner (13). The HLA-DRB1*03:01 allele is part of an extended risk haplotype that is in strong linkage disequilibrium with *C4A* copy number, and we have shown that lower *C4* copy number is associated with SLE and anti-SSA/SSB antibodies (14).

We hypothesized that a standardized procedure of quantifying DNA methylation changes at top DMCs in patients with SLE can be used to predict clinical presentation. The aim of the study was to investigate associations between a SLE methylation risk score (MRS) and clinical variables, and to delineate whether the phenotypic panorama defined by an elevated MRS can be distinguished from the clinical presentation linked to a high PRS. Further, we aimed to analyse the interrelations of the PRS and the MRS, across HLA-DRB1*03:01 positive and negative patients.

## Method

### Subjects and samples

The discovery cohort included 311 patients with SLE from Uppsala and Linköping University hospitals and a control group of 400 healthy blood donors visiting the Department of Transfusion Medicine, Uppsala University Hospital. The replication cohort comprised 175 patients from Karolinska University Hospital and 187 population controls (11). All patients fulfilled the American College of Rheumatology (ACR)-1982 classification criteria for SLE (15). Controls were matched for age and sex. Clinical data were collected from patient charts and included sex, age at sampling, ACR-82 criteria (15), autoantibodies (anti-SSA, anti-SSB, anti-RNP, anti-Sm and anti-dsDNA), disease activity measured as either SLE Disease Activity Index 2000 (SLEDAI-2K) (16) or Systemic Lupus Activity Measure (SLAM) (17), and medications at sampling. Simoa® was used to assess IFN-α2 concentration in serum (supplementary methods) (18).

### DNA methylation analysis

Patient and control whole blood DNA was subjected to Illumina HumanMethylation 450k BeadChip methylation array analysis, a microarray measuring DNA methylation levels at individual CpG sites on bisulfite-converted DNA and thereby generating methylation data of various regions across the genome (19). For detailed methodological information, see Imgenberg-Kreuz *et al.* 2018 (11). The DNA methylation analysis included analysis of methylation levels of 487 577 CpG sites in whole blood genomic DNA from patients and controls. Subsequently, signal intensity data were exported to Minfi R package for quality control (QC) and Subset-quantile Within Array Normalisation (SWAN), generating a post-QC dataset including 385 851 CpG sites.

### Methylation Analysis and development of the methylation risk score (MRS)

The MRS was calculated based on DNA methylation levels at previously published DMCs between patients with SLE and controls (Imgenberg-Kreuz *et al.* (11)). Significant DMCs with an effect size (β value) of ≥0.1, located at independent genetic loci, were selected rendering a total of 17 CpG sites, table 1.

**Table 1.**
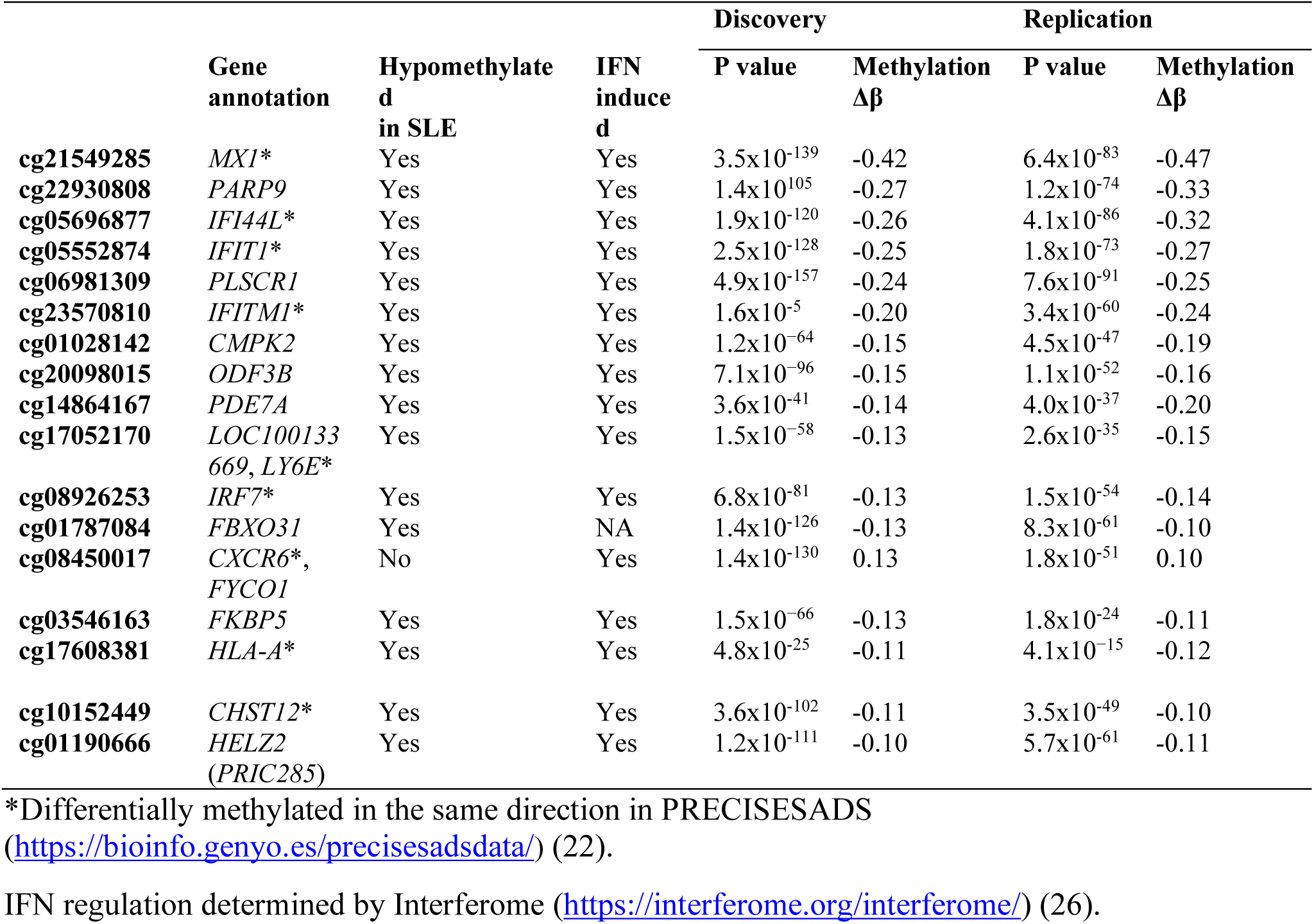
CpG sites included in the Methylation risk score (MRS)

The score was calculated according to the formula developed by Feng X *et al.* (20) and described for DNA methylation data by Björk A *et al.* (21). The mean and standard deviation (SD) of the methylation β value for each CpG site in the cohort-specific control group were used to achieve standardized values (Z-scores) for each individual:

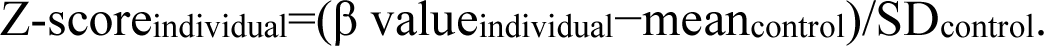

The final methylation scores were obtained through summation of the z-scores. Z-scores of CpG sites known to be hypomethylated in SLE were multiplied by −1. In total, 16 of the 17 CpG sites included in the MRS were hypomethylated. Next, genes annotated to the CpG sites included in the MRS were investigated for differential methylation in data from the PRECISESADS methylation browser (https://bioinfo.genyo.es/precisesadsdata/) (22).

Subsequently, a methylation score based on PRECISEADS-filtered DMCs, normalised against cohort-specific controls, was calculated for each individual.

### Genotyping and polygenic risk score development

In addition to the MRS, an SLE-PRS comprising 57 single nucleotide variants (SNVs), as described by Reid *et al.,* was analysed for associations with the same clinical subphenotypes in a combined cohort of patients from the discovery and replication datasets (n=486) (5).

Genotyping was performed using the Illumina 200K Immunochip SNP array, and the PRS was calculated by summation of the weighted risk alleles (supplementary methods). Lastly, genotype data for an HLA-DRB1*03:01 tag SNP (rs1269852) was extracted from the Immunochip dataset (23).

### Association with clinical variables

Associations between the MRS and clinical subphenotypes were first analysed separately in the discovery and the replication cohorts. Subsequently, a MRS normalised to all controls in the combined cohort was calculated for each patient, and analysed for associations with the PRS, the HLA-DRB1*03:01 tag SNP, and clinical parameters.

### Statistical analysis

Multivariable logistic regression models, with age and sex included as covariates, were applied to assess associations between the epigenetic and genetic scores and binary dependent variables. Continuous variables were analysed and compared between independent groups by Kruskal-Wallis rank sum test with post-hoc Dunn’s test, and by Wilcoxon rank-sum test when applicable. Linear regression models, adjusted for age and sex, were fitted to assess associations between continuous variables. Statistical analyses were conducted and plots were generated in R studio version 2024.09.1+394 (24).

### Ethics

Ethical approval was obtained for patients and controls (DNR 2009/013, 2016/155 and 2020-05065). All participants in the study provided their informed consent.

## Results

### MRS distribution in patients and controls

The methylation score included 17 CpG sites. The largest |Δβ| were observed for the CpG site annotated to *MX1*, followed by *PARP9* and *IFI44L* (table 1) (11). Patients with SLE exhibited significantly elevated and more widely distributed MRS values compared to controls in the discovery as well as the replication cohort (Fig. 1A, Supplementary Fig. S1). In the discovery cohort, male patients displayed significantly higher MRS scores compared to female patients with SLE (Supplementary Fig. S1). In the same cohort, older age at the time of sampling was associated with lower MRS in patients with SLE (B = (−0.39), p = 3.0 x 10^-5^), whereas a trend towards increased MRS levels was observed in healthy controls with older age at sampling (B = 0.036, p = 0.057) (Fig. 1B). The direction of association differed significantly between controls and patients in the discovery as well as the replication cohort (Fig. 1B and Supplementary Fig. S2).

**Fig. 1.**
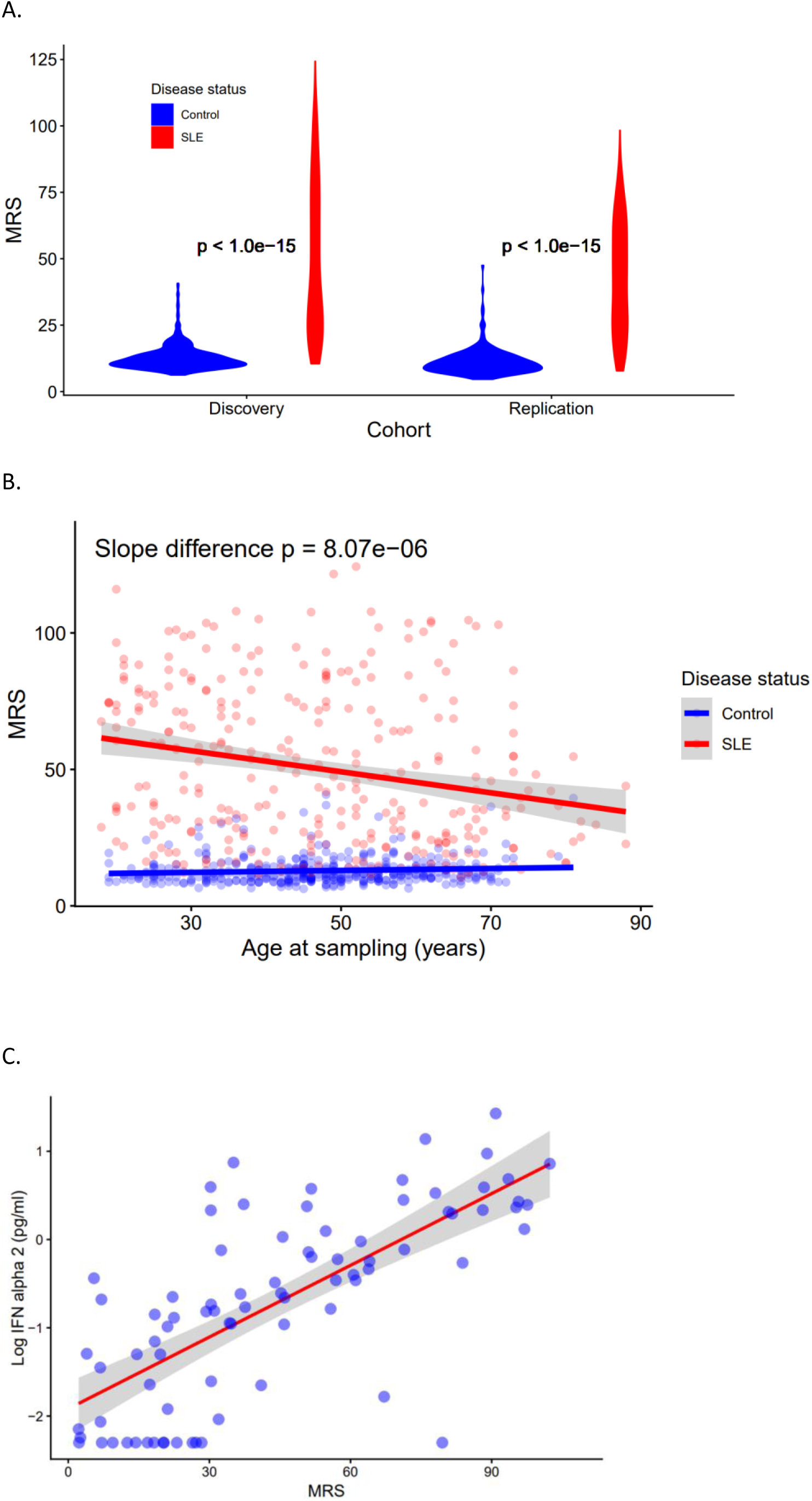
MRS distributions in SLE compared to controls, and across serum levels of IFN-α in a subset of patients. (A) Distribution of the MRS in patients and controls in the discovery and replication cohort. P-values, two-sided Mann-Whitney U test. (B) Scatter plot of the relationship between age at sampling and MRS levels across patients and controls in the discovery cohort. P value for difference between the two regression slopes determined by univariate analysis with disease status and age at sampling included with an interaction term. (C) Plot of correlation between the MRS and the common logarithm of Interferon alpha 2 (IFN-α2) concentration values (Simoa measurements) (18). MRS, Methylation Risk Score.

### MRS and levels of IFN-α

Since 16 of the 17 annotated genes were found to be IFN induced (Table 1), we examined whether the MRS associated with serum IFN-α2 levels as measured by Simoa (18). In a linear regression model adjusted for age at sampling and sex, higher MRS was associated with IFN-α2 levels (Exp(B)=1.03, p=1.04×10^−14^) (Fig. 1C). Overall, these findings demonstrate that the MRS integrates IFN-induced methylation changes, providing a quantitative measure that can differentiate patients and controls.

### Clinical subphenotypes and MRS

Levels of IFN-α have previously been demonstrated to link with various clinical manifestations of SLE (25). We therefore examined whether altered MRS predicted specific clinical subphenotypes, employing a discovery and replication cohort design. Higher MRS was associated with an increased risk of discoid lupus, immunologic and hematologic disorder in the discovery cohort, and with discoid rash and immunologic disorder in the replication cohort (Table 2). Higher MRS also associated with positivity for all the investigated autoantibodies in the discovery cohort, of which anti-SSA/SSB antibodies, anti-RNP antibodies and anti-Sm antibodies were confirmed in the replication data (table 2).

**Table 2.** Associations between the Methylation risk score (MRS) and clinical subphenotypes.

|  | Discovery |  |  | Replication |  |  |
| --- | --- | --- | --- | --- | --- | --- |
|  | N (%) | OR (95% CI) | p-value | N (%) | OR (95% CI) | p-value |
| ACR criteria* |  |  |  |  |  |  |
| Malar rash | 168 (54) | 1.00 (1.00–1.01) | $3.01 \times 10^{-1}$ | 88 (50.3) | 1.01 (0.99–1.02) | $3.38 \times 10^{-1}$ |
| Discoid lupus | 63 (20.3) | **1.02 (1.01–1.03)** | **$2.32 \times 10^{-3}$** | 26 (14.9) | **1.02 (1.00–1.05)** | **$4.50 \times 10^{-2}$** |
| Photosensitivity | 186 (59.8) | 1.01 (1.00–1.02) | $1.16 \times 10^{-1}$ | 119 (68) | 0.99 (0.97–1.01) | $1.81 \times 10^{-1}$ |
| Oral ulcers | 61 (19.6) | 1.00 (0.99–1.01) | $5.65 \times 10^{-1}$ | 46 (26.3) | 0.99 (0.98–1.01) | $5.25 \times 10^{-1}$ |
| Arthritis | 230 (74) | 0.99 (0.98–1.00) | $5.28 \times 10^{-2}$ | 146 (83.4) | 1.01 (0.99–1.03) | $3.44 \times 10^{-1}$ |
| Serositis | 129 (41.5) | 1.00 (1.00–1.01) | $4.14 \times 10^{-1}$ | 76 (43.4) | 1.01 (0.99–1.02) | $5.16 \times 10^{-1}$ |
| Nephritis | 90 (28.9) | 1.01 (1.00–1.01) | $2.71 \times 10^{-1}$ | 65 (37.1) | 1.01 (1.00–1.03) | $1.25 \times 10^{-1}$ |
| Neurologic | 18 (5.8) | 1.01 (1.00–1.03) | $9.00 \times 10^{-2}$ | 18 (10.3) | 1.01 (0.98–1.03) | $5.48 \times 10^{-1}$ |
| Hematologic | 186 (59.8) | **1.02 (1.01–1.03)** | **$2.02 \times 10^{-6}$** | 122 (69.7) | 1.02 (1.00–1.03) | $5.56 \times 10^{-2}$ |
| Immunologic | 187 (60.1) | **1.01 (1.00–1.02)** | **$1.06 \times 10^{-2}$** | 120 (68.6) | **1.02 (1.00–1.04)** | **$2.55 \times 10^{-2}$** |
| Anti-SSA | 143 (46.3) | **1.02 (1.01–1.03)** | **$1.01 \times 10^{-5}$** | 79 (45.1) | **1.06 (1.04–1.08)** | **$2.49 \times 10^{-8}$** |
| Anti-SSB | 85 (27.4) | **1.02 (1.01–1.03)** | **$2.67 \times 10^{-5}$** | 47 (26.9) | **1.04 (1.02–1.06)** | **$2.05 \times 10^{-4}$** |
| Anti-RNP | 106 (34.2) | **1.03 (1.02–1.04)** | **$1.25 \times 10^{-8}$** | 47 (26.9) | **1.03 (1.01–1.05)** | **$5.64 \times 10^{-4}$** |
| Anti-Sm | 43 (13.9) | **1.03 (1.01–1.04)** | **$2.15 \times 10^{-5}$** | 24 (13.7) | **1.02 (1.00–1.05)** | **$3.62 \times 10^{-2}$** |
| Anti-dsDNA | 180 (58.1) | **1.01 (1.00–1.02)** | **$2.32 \times 10^{-2}$** | 112 (64) | 1.01 (1.00–1.03) | $1.85 \times 10^{-1}$ |
| High disease act.<sup>†</sup> | 60 (19.3) | **1.02 (1.01–1.03)** | **$1.50 \times 10^{-4}$** | 78 (44.8) | 1.01 (0.99–1.02) | $3.52 \times 10^{-1}$ |
| Low compl<sup>‡</sup> | 129 (41.5) | **1.01 (1.01–1.02)** | **$8.62 \times 10^{-4}$** | n.a. | n.a. | n.a. |
| Treatment |  |  |  |  |  |  |
| Prednisolone | 192 (61.7) | **1.02 (1.01–1.03)** | **$4.21 \times 10^{-5}$** | 98 (56) | **1.02 (1.01–1.04)** | **$9.14 \times 10^{-3}$** |
| Antimalarials | 149 (47.9) | **0.99 (0.98–1.00)** | **$5.72 \times 10^{-3}$** | 67 (59.8) | 1.00 (0.98–1.02) | $9.06 \times 10^{-1}$ |
| Azathioprine | 50 (16.1) | 1.01 (1.00–1.02) | $1.03 \times 10^{-1}$ | 34 (25.4) | 1.00 (0.98–1.02) | $9.42 \times 10^{-1}$ |
| Mycophenolate | 34 (10.9) | 1.01 (1.00–1.03) | $5.39 \times 10^{-2}$ | 15 (9.9) | 0.99 (0.97–1.02) | $7.11 \times 10^{-1}$ |
| Methotrexate | 20 (6.4) | 1.02 (1.00–1.03) | $5.53 \times 10^{-2}$ | 8 (5.4) | 0.99 (0.96–1.03) | $6.22 \times 10^{-1}$ |

Multivariable logistic regression models. P-values adjusted for age at sampling and sex are presented with $p < 0.05$ marked in bold. Anti-SSA, anti-Sjögren's-syndrome-related antigen A antibodies; Anti-SSB, anti-Sjögren's syndrome type B antibodies; Anti-RNP, anti-ribonucleoprotein antibodies; Anti-Sm, anti-Smith antibodies; Anti-dsDNA, anti-double-stranded DNA.
\*Disease manifestations according to the American College of Rheumatology 82-criteria for SLE (ACR-82). <sup>†</sup>High disease activity defined as SLE activity measure (SLAM) $> 6$ (replication cohort) or SLE disease activity index (SLEDAI) $> 4$ (discovery cohort) (16, 17). <sup>‡</sup>Hypocomplementemia defined according to SLEDAI (16).

Furthermore, we found an association between increased MRS and higher disease activity and hypocomplementemia in the discovery cohort. However, the association with active disease did not reach statistical significance in the replication cohort, and complement data was not available (Table 2). Among the five treatment categories examined, higher MRS was significantly associated with prednisolone use in both cohorts, whereas lower MRS was linked to ongoing antimalarial treatment, albeit only in the discovery cohort (Table 2).

Given that the epigenetic loci included in the MRS were selected employing a CpG based procedure, we aimed to investigate whether a score based on CpG sites whose corresponding gene displayed altered methylation in PRECISESADS, associated with the similar subphenotypes as the original MRS. A subset (n=9) of the 17 CpG sites included in the MRS was annotated to a gene that was differentially methylated in the same direction in patients with SLE compared to controls, as reported in the PRECISESADS methylation browser (Supplementary Fig. S3) (22). The clinical predictability of the score was determined by analysis of associations with clinical variables in our discovery and replication cohort. The score was significantly associated with the similar clinical subphenotypes as the original MRS, including discoid rash, hematologic disorder, anti-SSA/SSB antibodies, anti-RNP antibodies and prednisolone use, and all these associations were significant in both the discovery and replication phase (Supplementary Table S1).

### MRS and PRS clinical and serological correlates

Since Reid *et al.* have previously demonstrated that a higher PRS was associated with a more severe disease including nephritis and anti-dsDNA antibodies, we proceeded to compare the clinical predictive properties of the MRS and the PRS (5). To facilitate this comparison, the discovery and replication cohorts were combined, rendering a total of 486 patients with SLE. In this combined cohort, the PRS was associated with nephritis and immunologic disorder, whereas an elevated MRS was linked to a significantly higher risk of discoid lupus, immunologic and hematologic disorder, and high disease activity. Higher MRS was significantly associated with positivity for all the investigated autoantibodies, with the highest level of significance observed for anti-SSA antibodies. In contrast, higher PRS only conferred increased risk of anti-dsDNA antibody positivity and associated with lower odds of anti-SSB antibodies (Supplementary Table S2). Patients with cumulative positivity for multiple autoantibodies exhibited significantly higher MRS levels (Fig. 2A). We further examined whether patients in the top 5% of MRS values exhibited increased risk of any specific clinical subphenotypes. This subgroup demonstrated higher prevalence of malar rash in addition to the previous observed associations (Supplementary Table S3). Overall, these findings suggest that the MRS captures a broader and to some extent different spectrum of clinical and serological features than the PRS, supporting its potential utility as a complementary biomarker in SLE.

**Fig. 2.**
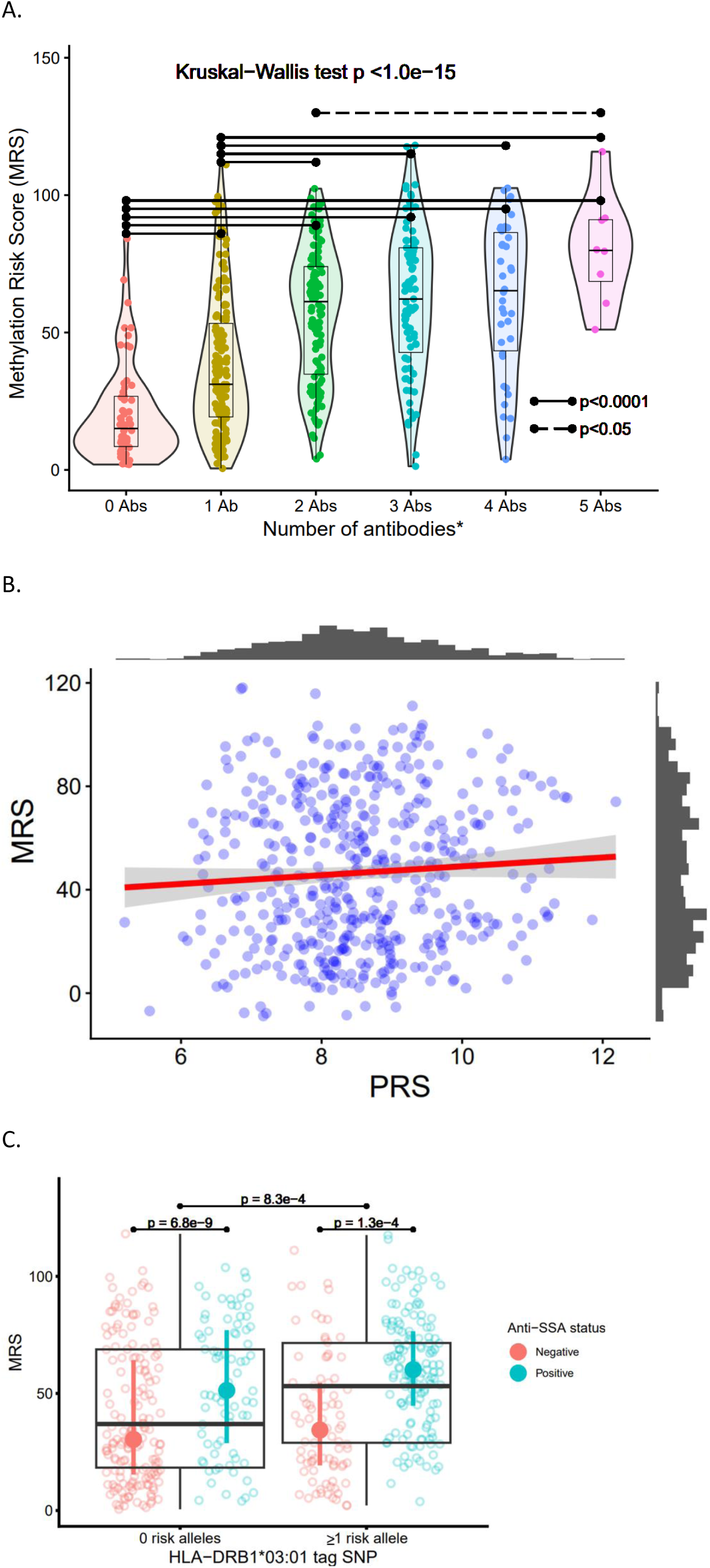
MRS associations with antibodies and the PRS, including stratification by HLA-DRB1*03:01 positivity. (A) Distribution of MRS values in patients with cumulative positivity for different autoantibodies. P values, Kruskal-Wallis rank sum test with post hoc Dunn’s test. P values displayed for all significant differences. (B) Scatter plot with fitted linear regression line of association between the PRS and the MRS. (C) Distribution of the MRS in patients that are non-carriers and carriers of the *HLA-DRB1*03:01* risk allele, stratified by anti-SSA antibody status. *Positivity ever for the autoantibodies anti-SSA, anti-SSB, anti-RNP, anti-dsDNA and anti-Sm. MRS, Methylation Risk Score; PRS, Polygenic Risk Score.

### Minimal overlap between the MRS and PRS

As only two shared associations were found when comparing MRS and PRS associations with clinical findings, we aimed to further explore the relationship between the two scores. In a linear regression model adjusted for age and sex, the scores were not significantly associated (B = 1.04, p = 0.35, R^2^ = 0.05) (Fig. 2B). Only one gene annotated to a CpG site in the MRS, *IRF7,* was represented in the PRS. The two scores also differed with respect to IFN regulation status. While 16 of the 17 DMCs included in the MRS were annotated to genes reported as IFN regulated in interferome, only 29 of the 53 genes covered by the PRS were IFN induced (Supplementary Fig. S5) (26). Altogether, the methylation pattern and interferon regulation at loci contributing to the PRS differ distinctly from genes captured by the MRS, consistent with the observed lack of association between the two scores.

### MRS and HLA-DRB1*03:01

With reference to the previously known association between the HLA-DRB1*03:01 risk allele and SSA antibodies and the observed link between the MRS and anti-SSA/SSB antibodies in the current study, we compared the MRS and an HLA-DRB1*03:01 tag SNP. Patients carrying at least one HLA-DRB1*03:01 risk allele compared to patients with a non-risk genotype displayed significantly higher MRS scores in the combined cohort (Fig. 2C). Further, patients positive for anti-SSA antibodies exhibited significantly higher MRS levels, which pertained to both risk allele carriers and non-carriers (Fig. 2C). In HLA-DRB1*03:01 negative patients, higher MRS conferred a significantly increased risk of nephritis (Supplementary Table S5). When comparing MRS association with the PRS across HLA-DRB1*03:01 carriers and non-carriers, the slope difference between the two regression lines was significant, with patients without the risk allele exhibiting a tendency towards higher MRS levels with higher polygenic risk (B = 2.9, p = 0.064, adjusted for age and sex) (Supplementary Fig. S6). In summary, we observed a novel association between an HLA-DRB1*03:01 tag SNP and higher MRS, that can be linked to specific SLE features and distinguished from PRS-associated subphenotypes.

## Discussion

In the present study, we investigated associations between an MRS and clinical subphenotypes in patients with SLE. Higher MRS associated with increased levels of IFN-α, discoid rash, multiple autoantibody seropositivity and HLA-DRB1*03:01 carriership. In contrast, a high non-HLA PRS was linked to greater prevalence of renal disease and lower odds of anti-SSB antibodies, suggesting partly different underlying pathogenic mechanisms. We therefore propose that an elevated MRS confers an epigenetic predisposition for enhanced type I IFN signaling, presenting clinically as distinct subphenotypes linked to HLA-DRB1*03:01.

We report that higher MRS, reflecting the cumulative contribution of the most differentially methylated CpG sites in SLE, correlated with higher levels of circulating IFN-α2. Notably, 16 of the 17 CpGs included in the score were hypomethylated, and most annotated genes were IFN-regulated, supporting a close link between the MRS and type I IFN signaling. With regard to methylation stability over time in SLE, Lanata *et al.* reported that a significantly smaller proportion of CpG sites within IFN-responsive genes remained stable longitudinally (53%) compared with CpG sites in non-IFN-related genes (87%)(27). Among the 20 CpG sites showing the greatest methylation changes over two years, four genes (*IFI44L*, *PARP9*, *IFITM1* and *MX1*) overlapped with those included in the MRS. These observations suggest that, although the MRS captures a biologically relevant IFN-associated methylation signature, some of its components may be dynamic. Thus, the overall stability of the MRS over time remains to be determined and warrants further longitudinal investigation.

Next, we investigated the relationship between MRS levels and age at the time of sampling across patients and controls, revealing divergent associations. Higher age at sampling was associated with lower MRS in patients, whereas controls displayed the opposite trend. This could be due to medication-associated epigenetic effects since medications can influence DNA methylation (28, 29). Alternatively, the epigenetic alterations observed in SLE may attenuate over time. Patients with a late-onset SLE often display a milder disease, which might be related to an age-associated decline in disease expression (30, 31). In line with this, a recent study reported age-associated epigenetic attenuation of IFN responsive genes in patients with SLE (32), consistent with the inverse relationship between age and MRS observed of the present study. Taken together, treatment-related epigenetic modulation and age-related remethylation represent possible, yet speculative, mechanisms underlying the age-related decrease in MRS levels in patients with SLE.

The subsequent analyses examined whether the MRS was associated with risk of specific disease manifestations and compared the findings with that of the PRS. Higher MRS, but not PRS, was associated with high disease activity and low complement, suggesting that DNA methylation changes are important determinants of disease activity. This is consistent with previous observations linking elevated IFN levels to active disease (33), and recent studies reporting DNA hypomethylation of genes including IFN-regulated genes in SLE to be associated with disease activity (34, 35). Higher MRS, but not PRS, also associated with increased risk of discoid rash, hematologic disorder and multiple autoantibodies including anti-SSA, anti-RNP and anti-Sm, while patients in the top 5% of MRS values additionally exhibit higher prevalence of malar rash. Indeed, high expression of type I IFN responsive genes have previously been linked to skin rash, leukocytopenia and autoantibodies (36–38). Hence, high MRS could reflect an epigenetic susceptibility to type I IFN-driven disease expression.

To confirm the relevance of the genes included in our MRS, we generated a methylation score based on nine of the 17 CpG sites corresponding to genes previously reported as differentially methylated in SLE. This filtered score showed associations with the same clinical subtypes as the full MRS, indicating that the included genomic regions robustly capture disease-relevant methylation changes. Notably, DMCs in three of the genes (*MX1*, *IFI44L* and *IFIT1)* have previously been linked to the same autoantibodies as the MRS (39). In contrast, a higher PRS conferred a significantly increased risk of renal disease and lower odds of anti-SSB antibody positivity, consistent with our previous findings that a high PRS predict a more severe disease and greater organ damage accrual (5). Altogether, the MRS captures reproducible epigenetic alterations in SLE that define distinct clinical phenotypes from those of the PRS, highlighting its potential as a complementary tool for patient stratification.

A higher MRS was also associated with the HLA-DRB1*03:01 haplotype. This finding, in conjunction with the MRS being associated with discoid rash, SSA/SSB antibodies and hematologic disorder, aligns with the previously described HLA-DRB1*03 dominated subgroup of SLE with higher prevalence of anti-SSA/SSB antibodies, skin rash and leukopenia (40). In addition, Lundtoft *et al.* reported an association between the HLA-DRB*03:01 haplotype, *C4A* copy number variation, and the presence of anti-SSA/SSB antibodies (14). Collectively, these findings suggest that increased autoantibody production in HLA-DRB1*03:01 positive patients may promote immune complex formation, thereby stimulating type I IFN production. It has been demonstrated that IFN-α, a type I IFN, can induce hypomethylation of genes including IFN responsive genes (41, 42). Hence, sustained IFN exposure could induce DNA methylation changes, further amplifying IFN signaling and contributing to disease expression including increased disease activity in patients with SLE.

Because genetic and epigenetic factors are known to interact in the pathogenesis of SLE, we investigated whether the MRS was associated with the PRS. Interestingly, there was no significant correlation. Only one of the genes connected to the DMCs included in the MRS, *IRF7*, overlapped with the 57 SNPs included in the PRS by Reid *et al.* (5). However, patients not carrying the HLA-DRB1*03:01 risk allele displayed a tendency towards increased MRS levels, and the regression slope differed significantly from that of HLA-DRB1*03:01 positive individuals. We propose that higher MRS, independently of the PRS, defines a clinical spectrum including HLA-DRB1*03:01 positivity, skin rash, antibody diversity and type I IFN signaling. Contrastingly, in patients lacking the strong pathogenetic contribution of the HLA-DRB1*03:01 haplotype, higher PRS may infer elevated MRS levels.

One major strength of the study stems from the sizes of the cohorts, including a total of 486 patients with SLE and 587 controls. The patients with SLE were well-characterised, and controls were matched for age and sex. Further, a discovery and replication cohort design was employed, increasing the reliability of the findings. The regression models were not adjusted for cell type proportions, which could be considered a limitation since changes in DNA methylation may be attributable to changes in cell type composition in patients compared to controls (28). However, bioinformatic methods can only partially account for this confounding factor, and changes in cell type proportions could itself be due to altered methylation in specific leukocyte subsets. The participants of the present study mainly comprised Caucasians, which is an important limitation. Studies including individuals of other ethnicities are warranted.

## Conclusion

We report that an SLE MRS, but not a PRS, associates with discoid rash, hematologic disorder, SSA-antibodies, high disease activity, low complement and elevated serum IFN-α. By contrast, high PRS is linked to nephritis and anti-dsDNA positivity. HLA-DRB1*03:01 positive patients displayed significantly increased MRS levels, and the lack of correlation between the MRS and the PRS suggests that epigenetic and polygenic risk contribute to SLE pathogenesis largely independently. These findings highlight the potential of MRS to identify an interferon-driven, clinically distinct SLE subset, complementing genetic risk scores in understanding disease heterogeneity.

## Acknowledgements

We thank Rezvan Kiani Dehkordi, Marianne Petersson and Karolina Tandre for collecting samples from patients and controls. DNA methylation analyses were performed at the SNP&SEQ Technology Platform at the National Genomics Infrastructure (NGI) hosted by Science for Life Laboratory in Uppsala, Sweden (www.genotyping.se; www.sequencing.se). We thank all study patients and blood donors.

## Data availability statement

Data are available upon reasonable request.

