## Supplemental figures and tables for "Methylation Risk Score Identifies an Interferon-Driven SLE Subset Distinct from Polygenic Risk"

**Supplementary figures and tables**

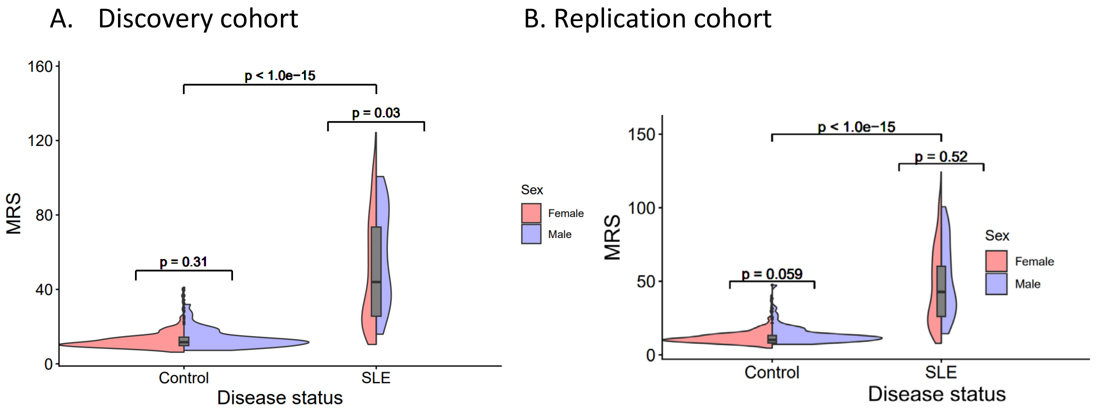

**Supplementary Figure S1.** Distribution of the MRS in patients compared to controls across male and female individuals. P-values, two-sided Mann Whitney-U test.

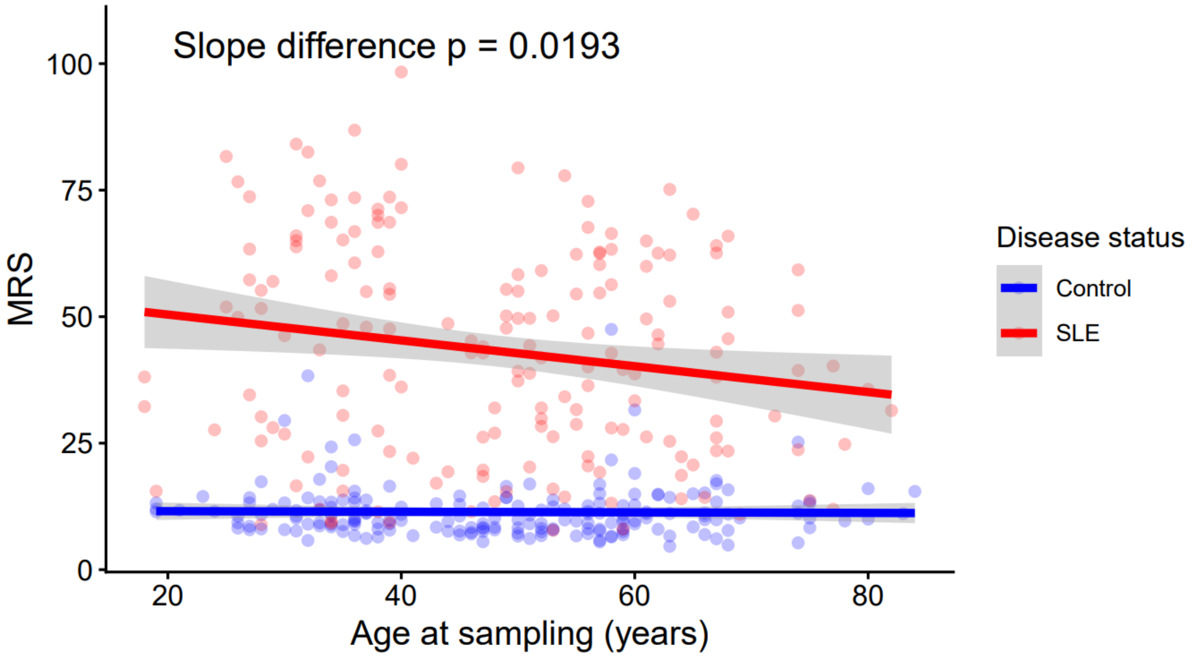

**Supplementary Figure S2.** Scatter plot of the relationship between age at sampling and MRS levels across patients and controls in the replication cohort. P value for difference between the two regression slopes determined by univariate analysis with disease status and age at sampling included with an interaction term.

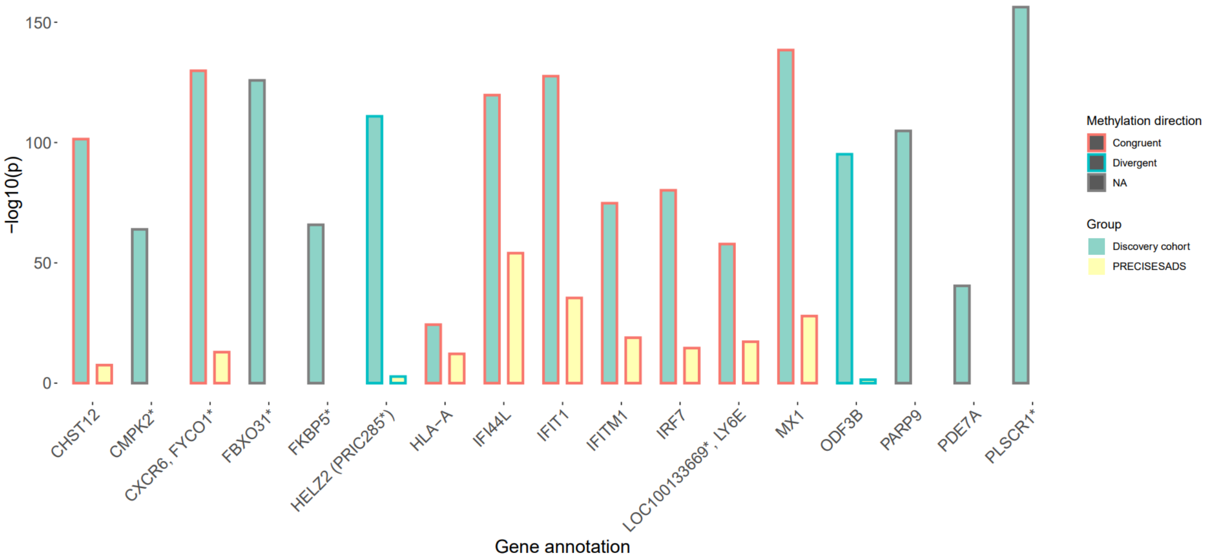

**Supplementary Figure S3.** Genes annotated to the CpG sites included in the MRS. Bars indicating level of statistical significance for the CpG sites included in the MRS, and for corresponding genes that were significantly differentially methylated in patients with SLE compared to controls in the PRECISESADS data set (1).

*No available methylation data in PRECISESADS for the specific gene.

**Supplementary Table S1.** Associations between the PRECISEADS-based MRS and clinical subphenotypes **(1)**.

| | **Discovery** | | | **Replication** | | |
|---|---|---|---|---|---|---|
| | **n** | **OR (95% CI)** | **p** | **n** | **OR (95% CI)** | **p** |
| **ACR criteria*** | | | | | | |
| **Malar rash** | 168 (54) | 1.01 (0.99–1.03) | $2.23\times10^{-1}$ | 88 (50.3) | 1.02 (0.99–1.06) | $1.55\times10^{-1}$ |
| **Discoid lupus** | 63 (20.3) | **1.04 (1.02–1.06)** | $\mathbf{6.49\times10^{-4}}$ | 26 (14.9) | **1.05 (1.00–1.11)** | $\mathbf{3.91\times10^{-2}}$ |
| **Photosensitivity** | 186 (59.8) | 1.01 (1.00–1.03) | $1.52\times10^{-1}$ | 119 (68) | 0.98 (0.95–1.02) | $3.78\times10^{-1}$ |
| **Oral ulcers** | 61 (19.6) | 1.00 (0.98–1.02) | $7.92\times10^{-1}$ | 46 (26.3) | 0.98 (0.95–1.02) | $3.50\times10^{-1}$ |
| **Arthritis** | 230 (74) | 0.98 (0.96–1.00) | $5.07\times10^{-2}$ | 146 (83.4) | 1.03 (0.98–1.07) | $2.57\times10^{-1}$ |
| **Serositis** | 129 (41.5) | 1.01 (0.99–1.03) | $3.91\times10^{-1}$ | 76 (43.4) | 1.02 (0.98–1.05) | $3.20\times10^{-1}$ |
| **Nephritis** | 90 (28.9) | 1.01 (0.99–1.03) | $1.72\times10^{-1}$ | 65 (37.1) | 1.02 (0.99–1.06) | $1.82\times10^{-1}$ |
| **Neurologic** | 18 (5.8) | 1.03 (0.99–1.07) | $1.14\times10^{-1}$ | 18 (10.3) | 1.01 (0.96–1.06) | $7.65\times10^{-1}$ |
| **Hematologic** | 186 (59.8) | **1.05 (1.03–1.07)** | $\mathbf{1.09\times10^{-6}}$ | 122 (69.7) | **1.04 (1.01–1.08)** | $\mathbf{2.22\times10^{-2}}$ |
| **Immunologic** | 187 (60.1) | **1.02 (1.01–1.04)** | $\mathbf{9.20\times10^{-3}}$ | 120 (68.6) | 1.03 (1.00–1.07) | $9.58\times10^{-2}$ |
| **Anti-SSA** | 143 (46.3) | **1.05 (1.03–1.07)** | $\mathbf{8.23\times10^{-7}}$ | 79 (45.1) | **1.13 (1.09–1.18)** | $\mathbf{1.25\times10^{-8}}$ |
| **Anti-SSB** | 85 (27.4) | **1.05 (1.03–1.07)** | $\mathbf{3.12\times10^{-6}}$ | 47 (26.9) | **1.09 (1.05–1.14)** | $\mathbf{7.91\times10^{-5}}$ |
| **Anti-RNP** | 106 (34.2) | **1.06 (1.04–1.08)** | $\mathbf{8.05\times10^{-9}}$ | 47 (26.9) | **1.07 (1.03–1.12)** | $\mathbf{4.53\times10^{-4}}$ |
| **Anti-Sm** | 43 (13.9) | **1.06 (1.03–1.09)** | $\mathbf{8.65\times10^{-5}}$ | 24 (13.7) | 1.04 (0.99–1.10) | $9.06\times10^{-2}$ |
| **Anti-dsDNA** | 180 (58.1) | **1.02 (1.00–1.04)** | $\mathbf{1.92\times10^{-2}}$ | 112 (64) | 1.02 (0.98–1.05) | $3.60\times10^{-1}$ |
| **High disease act.** † | 60 (19.3) | **1.04 (1.02–1.07)** | $\mathbf{1.32\times10^{-3}}$ | 78 (44.8) | 1.00 (0.97–1.04) | $7.81\times10^{-1}$ |
| **Low compl.**‡ | 129 (41.5) | **1.03 (1.01-1.05)** | $\mathbf{3.21\times10^{-3}}$ | n.a. | n.a. | n.a. |
| **Prednisolone** | 192 (61.7) | **1.03 (1.01–1.05)** | $\mathbf{6.64\times10^{-4}}$ | 98 (56) | **1.04 (1.01–1.07)** | $\mathbf{2.44\times10^{-2}}$ |
| **Antimalarials** | 149 (47.9) | **0.98 (0.96–0.99)** | $\mathbf{9.74\times10^{-3}}$ | 67 (59.8) | 1.00 (0.96–1.04) | $9.99\times10^{-1}$ |
| **Azathioprine** | 50 (16.1) | 1.02 (1.00–1.04) | $1.06\times10^{-1}$ | 34 (25.4) | 1.00 (0.96–1.04) | $9.30\times10^{-1}$ |
| **Mycophenolate** | 34 (10.9) | 1.02 (1.00–1.05) | $1.01\times10^{-1}$ | 15 (9.9) | 0.98 (0.92–1.04) | $5.28\times10^{-1}$ |
| **Methotrexate** | 20 (6.4) | 1.03 (1.00–1.06) | $8.00\times10^{-2}$ | 8 (5.4) | 0.98 (0.91–1.06) | $6.12\times10^{-1}$ |

Multivariable logistic regression models. P-values adjusted for age at sampling and sex are presented with $p<0.05$ marked in bold. Anti-SSA, anti-Sjögren's-syndrome-related antigen A antibodies; Anti-SSB, anti-Sjögren's syndrome type B antibodies; Anti-RNP, anti-ribonucleoprotein antibodies; Anti-Sm, anti-Smith antibodies; Anti-dsDNA, anti-double-stranded DNA.

*Disease manifestations according to the American College of Rheumatology 82-criteria for SLE (ACR-82). †High disease activity defined as SLE activity measure (SLAM) >6 (replication cohort) or SLE disease activity index (SLEDAI) >4 (discovery cohort) (2, 3). ‡Hypocomplementemia defined according to SLEDAI (2).

**Supplementary Table S2.** MRS and PRS associations with clinical subphenotypes. Combined cohort (n = 486).

| | | MRS | | PRS | |
|---|---|---|---|---|---|
| | **n** | **OR (95% CI)** | **p** | **OR (95% CI)** | **p** |
| **ACR-criteria*** | | | | | |
| **Malar rash** | 256 (52.7) | 1.00 (1.00–1.01) | $2.08\times10^{-1}$ | 1.02 (0.88–1.19) | $7.56\times10^{-1}$ |
| **Discoid lupus** | 89 (18.3) | **1.02 (1.01–1.02)** | $\mathbf{3.51\times10^{-4}}$ | 0.85 (0.69–1.04) | $1.14\times10^{-1}$ |
| **Photosensitivity** | 305 (62.8) | 1.00 (1.00–1.01) | $3.91\times10^{-1}$ | 0.91 (0.78–1.07) | $2.52\times10^{-1}$ |
| **Oral ulcers** | 107 (22) | 1.00 (0.99–1.01) | $9.64\times10^{-1}$ | 0.96 (0.80–1.16) | $6.68\times10^{-1}$ |
| **Arthritis** | 376 (77.4) | 1.00 (0.99–1.00) | $4.46\times10^{-1}$ | 0.99 (0.82–1.18) | $8.94\times10^{-1}$ |
| **Serositis** | 205 (42.2) | 1.00 (1.00–1.01) | $3.64\times10^{-1}$ | 0.88 (0.75–1.02) | $9.31\times10^{-2}$ |
| **Nephritis** | 155 (31.9) | 1.01 (1.00–1.01) | $5.71\times10^{-2}$ | **1.18 (1.00–1.40)** | $\mathbf{4.79\times10^{-2}}$ |
| **Neurologic** | 36 (7.4) | 1.01 (1.00–1.02) | $7.49\times10^{-2}$ | 1.12 (0.84–1.49) | $4.27\times10^{-1}$ |
| **Hematologic** | 308 (63.4) | **1.02 (1.01–1.03)** | $\mathbf{1.57\times10^{-7}}$ | 1.03 (0.88–1.20) | $7.34\times10^{-1}$ |
| **Immunologic** | 307 (63.2) | **1.01 (1.01–1.02)** | $\mathbf{1.39\times10^{-4}}$ | **1.40 (1.18–1.67)** | $\mathbf{1.12\times10^{-4}}$ |
| **Anti-SSA** | 222 (45.9) | **1.03 (1.02–1.03)** | $\mathbf{1.08\times10^{-12}}$ | 0.86 (0.73–1.00) | $5.16\times10^{-2}$ |
| **Anti-SSB** | 132 (27.2) | **1.02 (1.02–1.03)** | $\mathbf{2.77\times10^{-9}}$ | **0.81 (0.68–0.97)** | $\mathbf{2.21\times10^{-2}}$ |
| **Anti-RNP** | 153 (31.5) | **1.02 (1.02–1.03)** | $\mathbf{8.21\times10^{-11}}$ | 1.03 (0.88–1.22) | $6.91\times10^{-1}$ |
| **Anti-Sm** | 67 (13.8) | **1.02 (1.01–1.03)** | $\mathbf{4.94\times10^{-6}}$ | 1.05 (0.84–1.31) | $6.52\times10^{-1}$ |
| **Anti-dsDNA** | 292 (60.2) | **1.01 (1.00–1.02)** | $\mathbf{2.59\times10^{-3}}$ | **1.36 (1.16–1.61)** | $\mathbf{2.72\times10^{-4}}$ |
| **High disease act.** † | 138 (28.5) | **1.01 (1.01–1.02)** | $\mathbf{1.98\times10^{-4}}$ | 0.93 (0.78–1.10) | $3.96\times10^{-1}$ |
| **Low compl.**‡ | 129 (41.5) | **1.01 (1.00-1.02)** | $\mathbf{2.20\times10^{-3}}$ | 1.12 (0.92-1.37) | $2.60\times10^{-1}$ |

Multivariable logistic regression models. P-values adjusted for age at sampling and sex are presented with p<0.05 marked in bold. Anti-SSA, anti-Sjögren's-syndrome-related antigen A antibodies; Anti-SSB, anti-Sjögren's syndrome type B antibodies; Anti-RNP, anti-ribonucleoprotein antibodies; Anti-Sm, anti-Smith antibodies; Anti-dsDNA, anti-double-stranded DNA.

*Disease manifestations according to the American College of Rheumatology 82-criteria for SLE (ACR-82). †High disease activity defined as SLE activity measure (SLAM) >6 (replication cohort) or SLE disease activity index (SLEDAI) >4 (discovery cohort) (2, 3). ‡Hypocomplementemia defined according to SLEDAI (2).

**Supplementary Table S3.** Associations between exhibiting the 5% highest MRS values and clinical subphenotypes.

| ACR criteria* | n | OR (95% CI) | p |
|---|---|---|---|
| **Malar rash** | 486 | **2.86 (1.18–8.01)** | **$2.84\times10^{-2}$** |
| **Discoid lupus** | 486 | 2.00 (0.74–4.86) | $1.43\times10^{-1}$ |
| **Photosensitivity** | 486 | 1.24 (0.53–3.16) | $6.28\times10^{-1}$ |
| **Oral ulcers** | 486 | 1.61 (0.64–3.76) | $2.86\times10^{-1}$ |
| **Arthritis** | 486 | 0.50 (0.22–1.21) | $1.08\times10^{-1}$ |
| **Serositis** | 486 | 1.62 (0.71–3.69) | $2.46\times10^{-1}$ |
| **Nephritis** | 486 | 1.48 (0.61–3.43) | $3.65\times10^{-1}$ |
| **Neurologic** | 486 | 1.15 (0.18–4.13) | $8.58\times10^{-1}$ |
| **Hematologic** | 486 | **3.20 (1.19–11.11)** | **$3.63\times10^{-2}$** |
| **Immunologic** | 486 | 1.24 (0.53–3.15) | $6.31\times10^{-1}$ |
| **Anti-SSA** | 484 | 2.14 (0.94–5.16) | $7.55\times10^{-2}$ |
| **Anti-SSB** | 485 | 1.75 (0.74–3.99) | $1.87\times10^{-1}$ |
| **Anti-RNP** | 485 | **3.49 (1.54–8.23)** | **$3.02\times10^{-3}$** |
| **Anti-Sm** | 485 | **3.08 (1.19–7.38)** | **$1.42\times10^{-2}$** |
| **Anti-dsDNA** | 485 | 1.42 (0.61–3.58) | $4.33\times10^{-1}$ |
| **High disease act.** † | 485 | **2.32 (1.01–5.31)** | **$4.49\times10^{-2}$** |
| **Low compl.**‡ | 311 | **3.61 (1.35–10.79)** | **$1.37\times10^{-2}$** |

Multivariable logistic regression models. P-values adjusted for age at sampling and sex are presented with p<0.05 marked in bold. Anti-SSA, anti-Sjögren's-syndrome-related antigen A antibodies; Anti-SSB, anti-Sjögren's syndrome type B antibodies; Anti-RNP, anti-ribonucleoprotein antibodies; Anti-Sm, anti-Smith antibodies; Anti-dsDNA, anti-double-stranded DNA.

*Disease manifestations according to the American College of Rheumatology 82-criteria for SLE (ACR-82). †High disease activity defined as SLE activity measure (SLAM) >6 (replication cohort) or SLE disease activity index (SLEDAI) >4 (discovery cohort) (2, 3). ‡Hypocomplementemia defined according to SLEDAI (2).

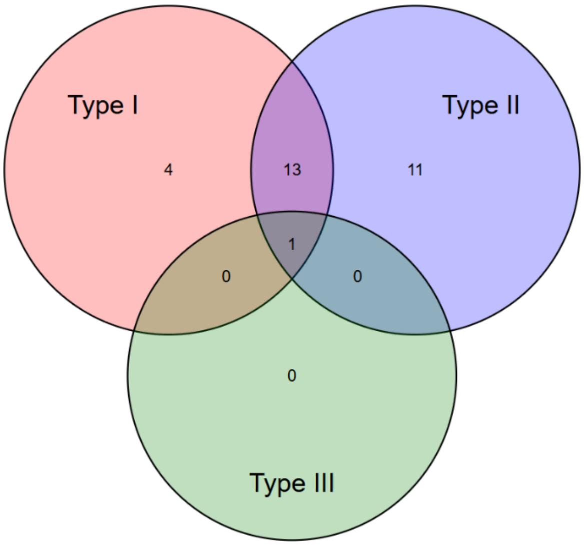

**Supplementary Figure S5.** Interferon regulation status of the genes annotated to the risk loci included in the PRS, as reported in the database Interferome (4).

**Supplementary Table S5.** Associations between the methylation risk score (MRS) and clinical variables in HLA-DRB1*03:01 negative and positive patients separately.

| | HLA-DRB1*03 :01 negative | | | HLA-DRB1*03 :01 positive | | |
|---|---|---|---|---|---|---|
| | **n** | **OR (95% CI)** | **p** | **n** | **OR (95% CI)** | **p** |
| **ACR-criteria*** | | | | | | |
| **Malar rash** | 258 | 1.01 (1.00–1.01) | $2.11\times10^{-1}$ | 228 | 1.00 (0.99–1.01) | $6.52\times10^{-1}$ |
| **Discoid lupus** | 258 | 1.02 (1.00–1.03) | $6.60\times10^{-3}$ | 228 | 1.02 (1.01–1.04) | $4.58\times10^{-3}$ |
| **Photosensitivity** | 258 | 1.00 (0.99–1.01) | $7.72\times10^{-1}$ | 228 | 1.01 (1.00–1.02) | $1.31\times10^{-1}$ |
| **Oral ulcers** | 258 | 1.00 (0.99–1.01) | $8.66\times10^{-1}$ | 228 | 1.00 (0.99–1.02) | $6.21\times10^{-1}$ |
| **Arthritis** | 258 | 1.00 (0.99–1.01) | $3.54\times10^{-1}$ | 228 | 1.00 (0.99–1.01) | $9.64\times10^{-1}$ |
| **Serositis** | 258 | 1.00 (0.99–1.01) | $7.51\times10^{-1}$ | 228 | 1.01 (1.00–1.02) | $1.47\times10^{-1}$ |
| **Nephritis** | 258 | 1.01 (1.00–1.02) | $7.24\times10^{-3}$ | 228 | 1.00 (0.99–1.01) | $8.36\times10^{-1}$ |
| **Neurologic** | 258 | 1.01 (0.99–1.02) | $2.57\times10^{-1}$ | 228 | 1.02 (1.00–1.04) | $7.99\times10^{-2}$ |
| **Hematologic** | 258 | 1.02 (1.01–1.03) | $1.20\times10^{-4}$ | 228 | 1.02 (1.01–1.03) | $7.51\times10^{-4}$ |
| **Immunologic** | 258 | 1.02 (1.01–1.03) | $2.80\times10^{-5}$ | 228 | 1.01 (1.00–1.02) | $2.77\times10^{-1}$ |
| **Anti-SSA** | 257 | 1.02 (1.01–1.03) | $2.38\times10^{-4}$ | 227 | 1.03 (1.02–1.05) | $4.88\times10^{-8}$ |
| **Anti-SSB** | 258 | 1.02 (1.01–1.04) | $5.61\times10^{-4}$ | 227 | 1.02 (1.01–1.03) | $8.17\times10^{-5}$ |
| **Anti-RNP** | 258 | 1.03 (1.02–1.04) | $1.94\times10^{-8}$ | 227 | 1.02 (1.01–1.03) | $6.92\times10^{-4}$ |
| **Anti-Sm** | 258 | 1.03 (1.02–1.04) | $1.80\times10^{-5}$ | 227 | 1.02 (1.00–1.03) | $4.66\times10^{-2}$ |
| **Anti-dsDNA** | 258 | 1.02 (1.01–1.03) | $1.26\times10^{-3}$ | 227 | 1.00 (0.99–1.01) | $4.17\times10^{-1}$ |
| **High disease act.** † | 257 | 1.02 (1.01–1.03) | $2.93\times10^{-4}$ | 228 | 1.01 (1.00–1.02) | $9.34\times10^{-2}$ |
| **Low compl.**‡ | 168 | 1.02 (1.01–1.03) | $4.85\times10^{-3}$ | 143 | 1.01 (1.00–1.02) | $1.13\times10^{-1}$ |

Multivariable logistic regression models. P-values adjusted for age at sampling and sex are presented with p<0.05 marked in bold. Anti-SSA, anti-Sjögren's-syndrome-related antigen A antibodies; Anti-SSB, anti-Sjögren's syndrome type B antibodies; Anti-RNP, anti-ribonucleoprotein antibodies; Anti-Sm, anti-Smith antibodies; Anti-dsDNA, anti-double-stranded DNA.

*Disease manifestations according to the American College of Rheumatology 82-criteria for SLE (ACR-82). †High disease activity defined as SLE activity measure (SLAM) >6 (replication cohort) or SLE disease activity index (SLEDAI) >4 (discovery cohort) (2, 3). ‡Hypocomplementemia defined according to SLEDAI (2)

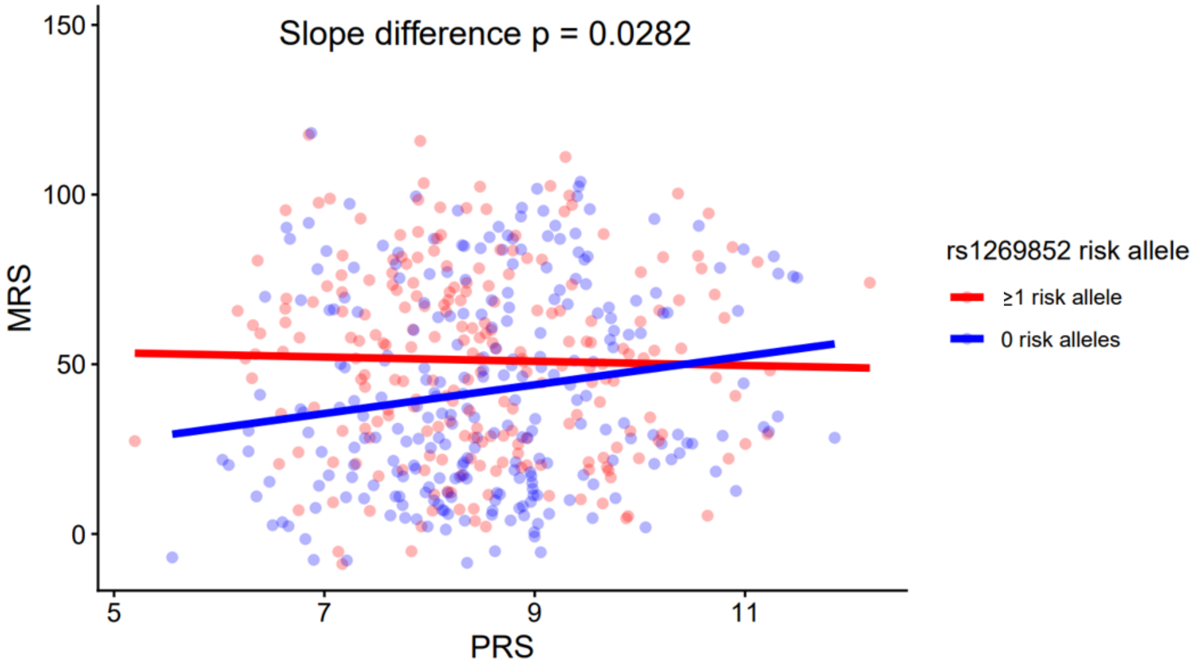

**Supplementary Figure S6.** Scatter plot with fitted linear regression lines of association between the PRS and the MRS across *HLA-DRB1*03:01* risk allele carriers and non-carriers. MRS, Methylation Risk Score; PRS, Polygenic Risk Score.
