## Supplemental methods for "Methylation Risk Score Identifies an Interferon-Driven SLE Subset Distinct from Polygenic Risk"

### Supplementary methods

**Genotyping and calculation of the PRS**

The Illumina 200K Immunochip SNP array was utilised for genotyping of the patients included in the study. Low-quality samples and variants were excluded using standard quality control (QC) protocols, and call rate cut-offs were set to 95% and 100% for samples and SNPs, respectively. A principal component analysis (PCA) of the 1000 Genomes Project data generated 5 principal components for the European cluster, and samples located outside 5 standard deviations (SD) in each of the principal components were excluded. Next, samples from subjects annotated as male with an inbreeding coefficient (F) close to zero and from subjects annotated as females with an F close to one were excluded, followed by exclusion of samples displaying abnormal autosomal heterozygosity rates as defined by >5 standard deviations from the mean Wright's F. The kinship coefficients based on identity-by-descent (IBD) was applied to examine cryptic relatedness. For each pair of samples related up to the second degree, one of the corresponding individuals was excluded. Finally, we omitted SNPs with a minor allele frequency (MAF) <1% or Hardy-Weinberg equilibrium (HWE) p-values < FDR 5% (based on controls only).

For calculation of the non-HLA PRS, SNPs previously shown to be associated with SLE at genome wide significance, that met the inclusion criteria described above, were included (1). Further, only subjects with complete genotyping for all 57 SNPs were included (2). In a cohort of 1001 patients and 2802 controls, the natural logarithm of the OR for SLE susceptibility was determined for each SNP. A PRS was assigned to each patient by multiplying the logarithm of the OR for each SNP by the number of risk alleles in the specific individual. HLA-SNPs shown to be associated with SLE at genome wide significance were filtered for independent signals, with variants with the lowest odds ratio for SLE for SNPs in linkage disequilibrium ($r^2>0.2$) being excluded. A SNP previously described as a tag SNP for HLA-DRB1*03:01, rs1269852, was selected (3).

**Measurement of serum IFN-α2**

Serum IFN- α2 concentrations were analysed using Simoa®, a Single Molecule Array platform on the SR-X™ Biomarker Detection System (Quanterix Corp. Billerica, Ma, USA) (4). All serum samples were subjected to 1:4 dilution. The lower limit of detection (LLOD) was 0.003 pg/ml, and the lower limit of quantification (LLOQ) was 0.05 pg/ml. Due to heteroscedasticity in the residuals when fitting the linear regression model, logarithmized IFN-α2 concentration values were analysed.

1. Chen L, Morris DL, Vyse TJ. Genetic advances in systemic lupus erythematosus: an update. Curr Opin Rheumatol. 2017;29(5):423-33.
2. Reid S, Alexsson A, Frodlund M, Morris D, Sandling JK, Bolin K, et al. High genetic risk score is associated with early disease onset, damage accrual and decreased survival in systemic lupus erythematosus. Ann Rheum Dis. 2020;79(3):363-9.
3. Hedenstedt A, Reid S, Sayadi A, Eloranta ML, Skoglund E, Bolin K, et al. B cell polygenic risk scores associate with anti-dsDNA antibodies and nephritis in systemic lupus erythematosus. Lupus Sci Med. 2023;10(2).
4. Rodero MP, Decalf J, Bondet V, Hunt D, Rice GI, Werneke S, et al. Detection of interferon alpha protein reveals differential levels and cellular sources in disease. J Exp Med. 2017;214(5):1547-55.
